# Prospective genomic surveillance and characterization of respiratory syncytial virus in Minnesota, USA – July 2023 to February 2024

**DOI:** 10.1101/2024.07.10.24310215

**Authors:** Daniel Evans, Henry Kunerth, Erica Mumm, Sarah Namugenyi, Matthew Plumb, Sarah Bistodeau, Scott A. Cunningham, Bryan Schmitt, Karen Martin, Katherine Como-Sabetti, Ruth Lynfield, Xiong Wang

## Abstract

We expanded Minnesota’s viral genomic surveillance program to include respiratory syncytial virus (RSV). We performed whole-genome sequencing of 575 specimens collected at Minnesota healthcare facilities from July 2023 to February 2024. Subtypes A and B exhibited differences in their genomic landscapes, and we identified 23 clusters of genetically identical genomes.

## INTRODUCTION

Respiratory syncytial virus (RSV) is a major respiratory pathogen, with increased risk of severe infections among infants and young children, elderly persons, and persons with underlying health conditions, including immunocompromise (1). Whole-genome sequencing (WGS) has been applied for retrospective RSV surveillance and outbreak investigation in the United States (2), but it has not yet been documented as a tool for prospective surveillance. Several genetic typing schemes have been developed for more granular characterization of RSV (3-4), but their use in genomic epidemiology has not been thoroughly evaluated. Here, we report preliminary findings from our recently established genomic surveillance of RSV in the state of Minnesota, one of the first such initiatives conducted in the United States.

## THE STUDY

From July 2023 through February 2024, we performed WGS on positive RSV respiratory specimens from outpatients and inpatients collected from eleven healthcare facilities in Minnesota. Institutional review board approval was not necessary, since this study was conducted as a component of public health surveillance subject to Minnesota Reporting Rules. Each specimen was submitted with limited patient data (name, sex assigned at birth, date of birth, date of specimen collection, and outpatient or inpatient status).

We amplified genomes from specimens using tiled amplicon-based primers (5) and sequenced genomes using the GridION platform (Oxford Nanopore Technologies). We performed genome assembly, quality control, and viral subtyping using the nf-core Viralrecon pipeline (6). We used Nextclade software to subtype genomes based on G-protein genotyping and whole-genome lineage typing schemes described in Goya et al, 2020 and Goya et al, 2024 (3-4, 7). We successfully sequenced 575 RSV genomes, of which 287 (49.9%) were classified as subtype A and 288 (50.1%) as subtype B (Table 1). All RSV-A and RSV-B genomes belonged to single G-protein genotypes – GA.2.3.5 and GB.5.0.5a, respectively. Nearly all RSV-A genomes (n = 285, 99.3%) were distributed among four whole-genome lineages: A.D.1 (n = 31, 10.8%), A.D.3 (n = 53, 18.5%), A.D.5 (n = 109, 38.0%), and A.D.5.2 (n = 92, 32.1%). By contrast, most RSV-B genomes (n = 274, 95.1%) belonged to a single lineage: B.D.E.1.

**TABLE 1:**
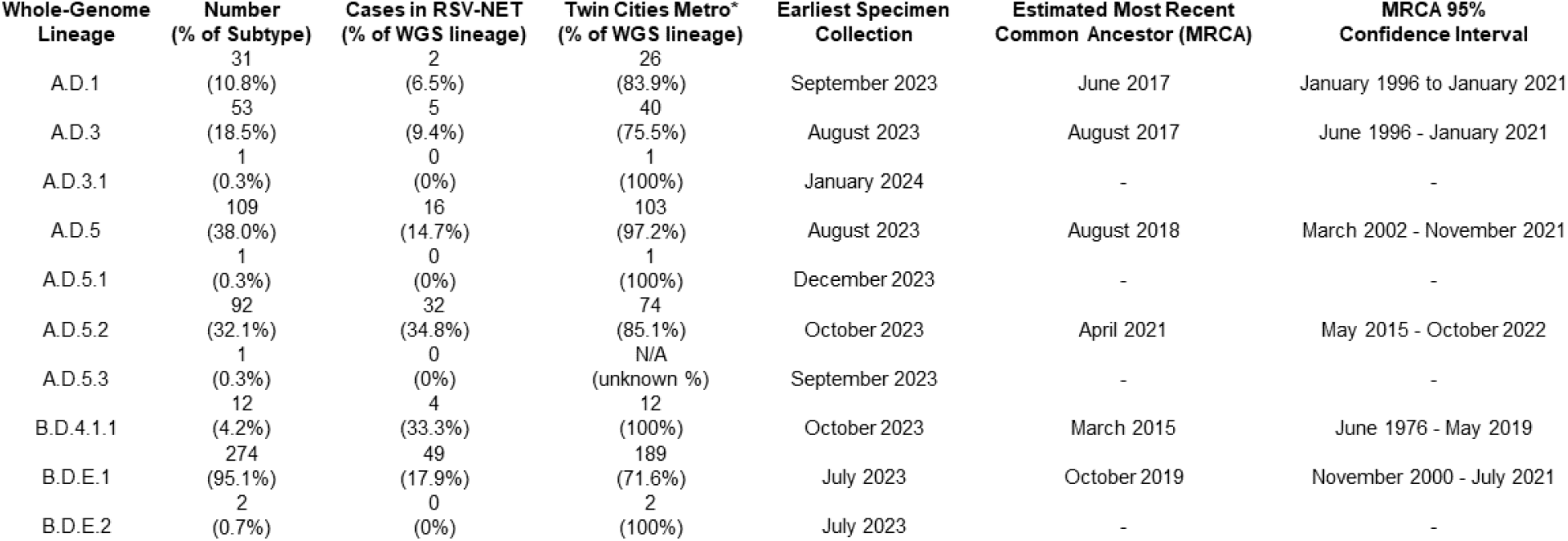
Summary of WGS results for RSV-A and RSV-B specimens. Counts and percentages exclude out-of-state cases and cases of unknown residence.

| Whole-Genome Lineage | Number<br>(% of Subtype) | Cases in RSV-NET<br>(% of WGS lineage) | Twin Cities Metro*<br>(% of WGS lineage) | Earliest Specimen<br>Collection | Estimated Most Recent<br>Common Ancestor (MRCA) | MRCA 95%<br>Confidence Interval |
| --- | --- | --- | --- | --- | --- | --- |
| A.D.1 | 31<br>(10.8%) | 2<br>(6.5%) | 26<br>(83.9%) | September 2023 | June 2017 | January 1996 to January 2021 |
| A.D.3 | 53<br>(18.5%) | 5<br>(9.4%) | 40<br>(75.5%) | August 2023 | August 2017 | June 1996 - January 2021 |
| A.D.3.1 | 1<br>(0.3%) | 0<br>(0%) | 1<br>(100%) | January 2024 | - | - |
| A.D.5 | 109<br>(38.0%) | 16<br>(14.7%) | 103<br>(97.2%) | August 2023 | August 2018 | March 2002 - November 2021 |
| A.D.5.1 | 1<br>(0.3%) | 0<br>(0%) | 1<br>(100%) | December 2023 | - | - |
| A.D.5.2 | 92<br>(32.1%) | 32<br>(34.8%) | 74<br>(85.1%) | October 2023 | April 2021 | May 2015 - October 2022 |
| A.D.5.3 | 1<br>(0.3%) | 0<br>(0%) | N/A<br>(unknown %) | September 2023 | - | - |
| B.D.4.1.1 | 12<br>(4.2%) | 4<br>(33.3%) | 12<br>(100%) | October 2023 | March 2015 | June 1976 - May 2019 |
| B.D.E.1 | 274<br>(95.1%) | 49<br>(17.9%) | 189<br>(71.6%) | July 2023 | October 2019 | November 2000 - July 2021 |
| B.D.E.2 | 2<br>(0.7%) | 0<br>(0%) | 2<br>(100%) | July 2023 | - | - |

We constructed phylogenetic trees using Nextstrain’s Augur pipeline v24.3.0 (Figures 1 and 2) (8-9). Briefly, we aligned viral genome assemblies to Nextstrain’s default reference sequences (hRSV/A/England/397/2017 and hRSV/B/Australia/VIC-RCH056/2019) using MAFFT v7.526 (10), and then constructed and refined distance- and time-scaled trees from those alignments using IQTree v2.3.3 and TimeTree v0.11.3 (11-12). Comparisons of tree architecture showed greater genetic diversity among RSV-A genomes (mean pairwise p-distance 0.0114) than among RSV-B genomes (mean pairwise p-distance 0.0045) (13). Time-scaled phylogenetic analysis estimated the divergence of whole-genome lineages to have occurred between 2 and 8 years before the earliest collection of a genome of that lineage (Table 1). These estimates were imprecise – likely due to the geographically and temporally localized sampling window – but 95% confidence intervals excluded dates that were more recent than 1 to 5 years before earliest specimen collection. Examining tree structure showed that 69.0% of genomes (RSV-A: n = 275, 95.8%; RSV-B: n = 122, 42.4%) were grouped in clades of at least 3 sequences which, with at least 95% confidence, diverged within 6 months of when their first viral specimens were collected.

**FIGURE 1:**
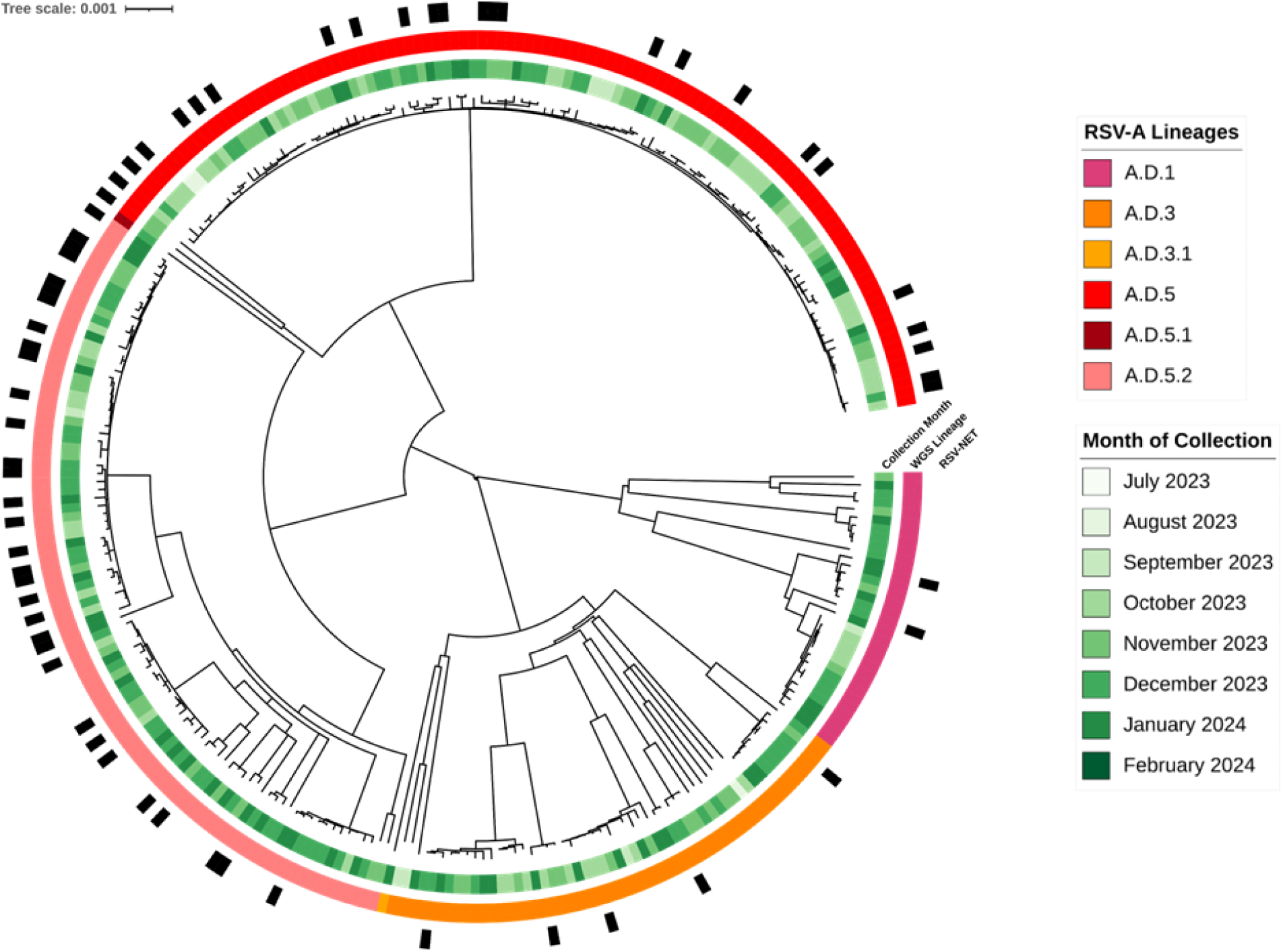
Midpoint-rooted phylogenetic trees of RSV-A genomes. Annotations denote the following, from innermost to outermost rings: month of specimen collection, whole-genome lineage type (4), and documentation of the case in the RSV-NET database of RSV-associated hospitalizations and deaths (15).

**FIGURE 2:**
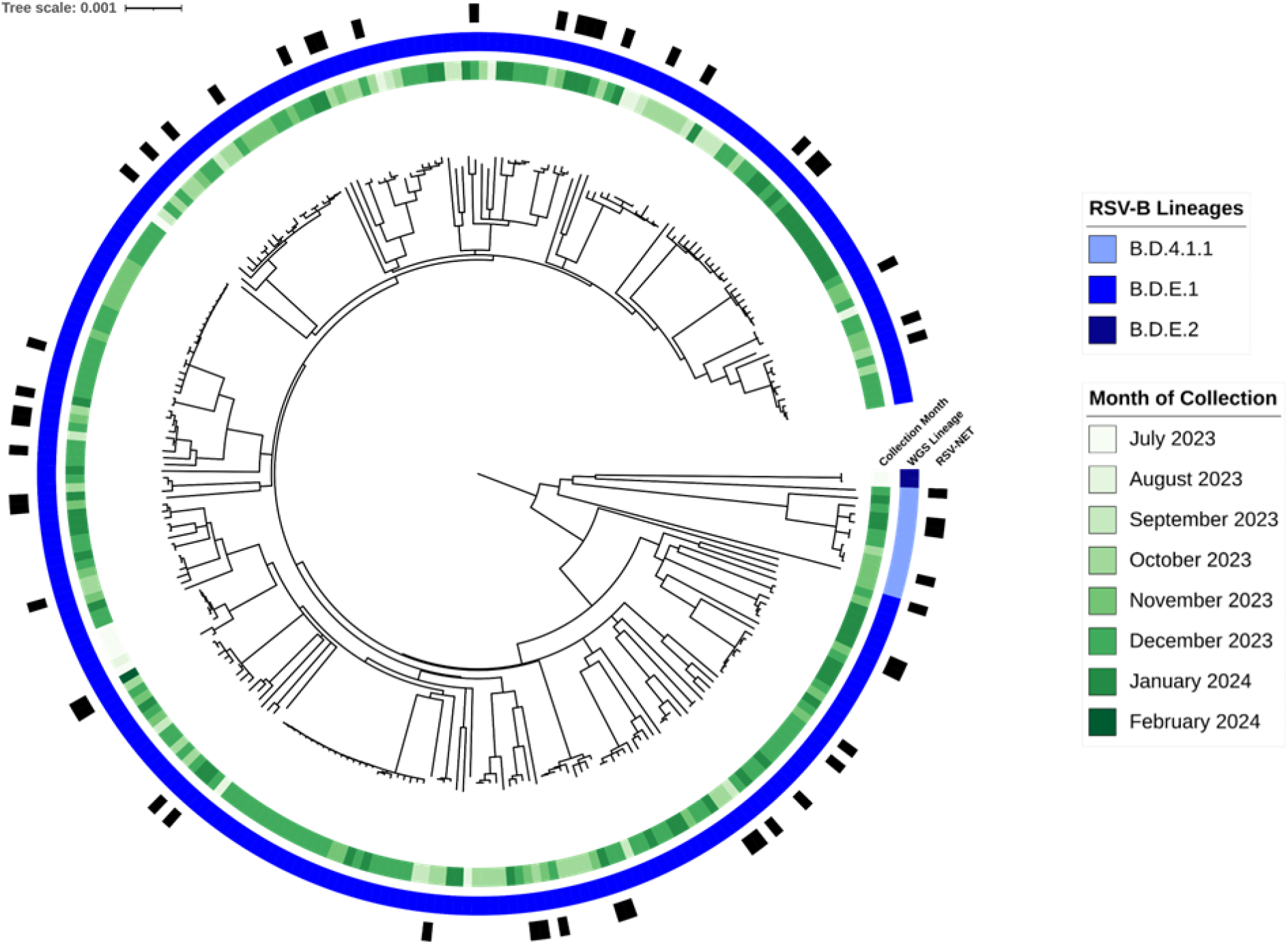
Midpoint-rooted phylogenetic trees of RSV-B genomes. Annotations denote the following, from innermost to outermost rings: month of specimen collection, whole-genome lineage type (4), and documentation of the case in the RSV-NET database of RSV-associated hospitalizations and deaths (15).

We identified single nucleotide polymorphisms (SNPs) from whole-genome alignments using SNP-dists v0.8.2 (14). Pairwise comparisons showed that 32.3% of genomes were identical to at least one other genome at 0 SNPs (RSV-A: n = 97, 33.7%; RSV-B: n = 89, 30.9%) and 53.0% within 1 SNP (RSV-A: n = 155, 54.0%; RSV-B: n = 150, 52.1%). We found 23 clusters of at least 3 genomes with shared nucleotide identity at 0 SNPs (RSV-A: n = 14, RSV-B; n = 9). These clusters included 19.5% of all genomes (RSV-A: n = 59, 20.6%; RSV-B: n = 53, 18.4%). SNP analyses further contrasted the population structures of subtypes A and B, as cumulative distribution functions of pairwise SNP calls within each subtype differed significantly (Kolmogorov-Smirnov D-statistic = 0.658, p < 0.001).

For all genomes, we cross-referenced patient name and date of birth with data from the Respiratory Syncytial Virus Hospitalization Surveillance Network (RSV-NET), a component of CDC’s Emerging Infections Program (EIP) conducting population-based surveillance of RSV-associated hospitalizations (Table 2, Figure 1) (15). Among 531 genomes collected between October 2023 and January 2024 – the peak months of specimen collection – 116 (21.8%) were from RSV-NET cases (RSV-A: n = 64, 55.2%; RSV-B: n = 52, 44.8%). These represented 6.3% of all hospitalized cases during that period. Nine of the 23 clusters (39.1%) included at least one RSV-NET case (RSV-A: n = 7, 50.0%; RSV-B: n = 2, 22.2%), and 13.4% of clustered cases were in RSV-NET (RSV-A: n = 12, 20.3%; RSV-B: n = 3, 5.7%).

**TABLE 2:** Comparison of Minnesota RSV cases with WGS data to hospitalized and/or deceased cases documented in RSV-NET, October 2023 – January 2024.

|  | **RSV-NET Cases,**<br>**Oct 2023 - Jan 2024** | **RSV Cases with WGS Data,**<br>**Oct 2023 - Jan 2024** | **RSV-NET Cases with WGS Data,**<br>**Oct 2023 - Jan 2024** |
| --- | --- | --- | --- |
| **Count** | 1847 | 524 | 116 |
| **Median Age** | 2 years | 1.7 years | 1 year |
| **% Female** | 51.1% | 49.2% | 51.7% |
| **% White** | 63.6% | 45.2% | 53.5% |
| **% metro** | 65.0% | 75.2% | 87.1% |
| **% of RSV-NET** | - | - | 6.30% |
| **% of WGS** | - | - | 20.2% |

The cohort of sequenced RSV-NET cases was not representative of the full population of RSV-NET cases (Table 2). While sequenced and all RSV-NET cases did not differ significantly by sex assigned at birth, sequenced cases were younger (median age 1 year vs. 2 years, p = 0.0193) and included a smaller proportion of cases of White versus non-White race (51.7% vs 63.6%, p = 0.0014). Sequenced RSV-NET cases were much more likely to live within the Minneapolis-St. Paul metropolitan area (87.1% vs. 65.0%, p < 0.001). These differences reflect the disproportionately high concentration of specimens submitted for sequencing from healthcare facilities in the area.

## CONCLUSIONS

This study summarizes early findings from a new statewide genomic surveillance program of RSV infections in Minnesota, USA. Our results show that applying WGS for RSV surveillance can yield insights into viral circulation and population dynamics. Prospective sequencing revealed differing genomic landscapes of subtypes A and B and contextualized the genetic diversity of RSV-A and RSV-B within the state. The detection of genetic clusters within the viral population shows the potential use of WGS for detection and investigation of RSV outbreaks.

In addition to surveying viral diversity, prospective genomic surveillance of RSV could also yield useful insights into mutations, clades, or lineages associated with increased transmissibility, greater virulence, or evolution of vaccine resistance (2-4). However, our study’s limitation of skewed convenience sampling for – and inconsistent quantities of epidemiological data linked to – specimens submitted for WGS prevented us from drawing reliable conclusions on these potential associations. Our future work will include expanding the collection of these data and more targeted collection of specimens, to improve representativeness in sampling design such that these questions can be investigated.

Daniel Evans is a Genomic Epidemiologist with the Minnesota Department of Health. His work focuses on developing, implementing, and optimizing the use of genomics for pathogen surveillance.

## Data Availability

All genome sequencing reads from this study are publicly available on NCBI under BioProject number PRJNA1048457. De-identified epidemiological data in the present study may be requested, subject to HIPAA and Minnesota state law.

https://www.ncbi.nlm.nih.gov/bioproject/?term=PRJNA1048457

## ACKNOWLEDGEMENTS

We thank the following healthcare facilities for submitting specimens: Carris Rice, CCM Health-Montevideo, Children’s Minnesota, Fairview Ridges, Fairview Southdale, Grand Itasca, Hennepin County Medical Center, Maple Grove Hospital, the Hennepin County medical examiner’s office, Minnesota State University-Mankato clinic, Ridgeview-Waconia, and Riverview-Crookston.

We thank the CDC RSV-NET program and the Emerging Infections Program (EIP) for their support of sentinel RSV surveillance in Minnesota, as well as Maloney et al for their support with laboratory methods.

This project was funded by CDC Epidemiology and Laboratory Capacity grant NU50CK000508 and Pathogen Genomics Centers of Excellence grant NU50CK000628.

All sequencing reads from this study are publicly available on NCBI under BioProject number PRJNA1048457.

## Notes

### Competing Interest Statement

The authors have declared no competing interest.

### Author Declarations

The Health Protection Bureau of the Minnesota Department of Health waived ethical approval for this study, since all data collection and analysis were conducted as a component of public health surveillance activities subject to Minnesota Reporting Rules.

